# A genomic-led strategy to anticipate drug safety effects

**DOI:** 10.1101/2025.09.11.25335448

**Authors:** Brian R Ferolito, Andrea RVR Horimoto, Kai Gravel-Pucillo, Daniel J Golden, Hesam Dashti, Claudia Giambartolomei, Danielle Rasooly, Rachael Matty, Liam Gaziano, Yakov Tsepilov, Lauren Costa, Nicole Kosik, Harris Ioannidis, Mohd Karim, Giovanna Winicki, Fiona Hunter, Claudia Langenberg, John C Whittaker, Million Veteran Program, Tianxi Cai, Gina M Peloso, Barbara Zdrazil, Maya Ghoussaini, Andrew R Leach, Sumitra Muralidhar, Ines A Smit, Juan P Casas, J Michael Gaziano, Kelly Cho, Alexandre C Pereira

**Affiliations:** Million Veteran Program (MVP) Coordinating Center, Veterans Affairs Healthcare System, 2 Avenue de Lafayette, Boston, 02111, MA, USA; Department of Genetics, University of North Carolina School of Medicine, 120 Mason Farm Road, Chapel Hill, 27599, NC, USA; Division of Aging, Brigham and Women’s Hospital and Harvard Medical School, 75 Francis St, Boston, 02115, MA, USA; Broad Institute of MIT and Harvard, 415 Main St., Cambridge, 02142, MA, USA; Health Data Science Centre, Human Technopole, V.le Rita Levi-Montalcini, 1, Milan, 20157, Italy; Centre for Human Technologies (CHT), Insituto Italiano di Tecnologia, Via Enrico Melen 83, Building B, 7th floor, Genova, 16152, Italy; BHF Cardiovascular Epidemiology Unit, University of Cambridge, Cambridge, CB2 0BB, UK; Open Targets, Hinxton, CB10 1SD, UK; European Molecular Biology Laboratory, European Bioinformatics Institute, Hinxton, CB10 1SD, UK; Genomic Discovery, Variant Bio, Seattle, 98109, WA, USA; Precision Healthcare University Research Institute, Queen Mary University of London, London, UK; Computational Medicine, Berlin Institue of Health at Charit′e – Universitätsmedizin, Berlin, Germany; MRC Epidemiology Unit, University of Cambridge, Cambridge, CB2 0SR, UK; MRC Biostatistics Unit, University of Cambridge, Cambridge, CB2 0SR, UK; Department of Biomedical Informatics, Harvard Medical School, Boston, 02115, MA, USA; Department of Biostatistics, Boston University School of Public Health, 801 Massachusetts Ave Crosstown Center, Boston, 02115, MA, USA; Regeneron Genetics Center, Regeneron Pharmaceuticals, Tarrytown, NY, 10591, USA; LifeArc, Accelerator Building, Open Innovation Campus, Stevenage, SG1 2FX, UK; Office of Research and Development, Department of Veterans Affairs, Washington, 20420, DC, USA; Biomarker Development/Translational Medicine, Novartis Institutes for Biomedical Research, 250 Massachusetts Ave, Cambridge, 02139, MA,USA; Department of Medicine, Harvard Medical School, Boston, 02115, MA, USA

## Abstract

Safety-related issues account for approximately 28% of failures in new drug discovery programs. On top of that, many are discovered during post-marketing surveillance, significantly limiting drug utility and application. To proactively address these concerns, we developed a genetics-led strategy leveraging Mendelian Randomization (MR) across large-scale genetic datasets from the Million Veteran Program, FinnGen, and UK Biobank. By mapping genetic variants associated with gene expression and protein abundance to 1,449 harmonized human phenotypes, we systematically identified potential adverse drug reactions (ADR). Our extensive MR analysis, encompassing 16,915 protein-coding genes, demonstrated the capacity to predict hundreds of known ADR for approved medications, with approximately 40% corroborated by FDA Adverse Event Reporting System (FAERS) data. Additionally, we found significant enrichment of identified gene-mechanism pairs in clinical trials terminated early due to safety concerns, highlighting the clinical utility of genetics-informed safety prediction. Notably, immune-related pathways were prominently associated with ADR, indicating particular sensitivity within immune modulation targets. Our comprehensive atlas, integrating genetic evidence with pharmacological mechanisms, provides a robust predictive framework for anticipating drug safety, potentially enhancing decision-making in drug development and pharmacovigilance.

## Introduction

Safety-related issues are responsible for approximately 24% of new drug discovery program failures, and even among drugs that reach approval, unexpected adverse drug reactions (ADRs) often surface only after widespread clinical use(*1, 2*). The development of a framework to predict adverse drug reactions (ADR) could substantially increase the success rate of drug development programs as well as the identification and reporting of ADR for novel and existing drugs(*3*). In this context, even partially predictive models for ADRs are extremely valuable because they can alter the course of drug development, regulatory review, and post-marketing monitoring. During early drug development, a partially predictive model could flag compounds at risk of hepatotoxicity, prompting additional toxicity assays or careful patient selection that could avert failures(*4*). In regulatory review, such models could prioritize drugs for enhanced risk management plans, as seen with clozapine, where modest genetic predictors help guide monitoring strategies for rare but serious blood disorders(*5*). Post-marketing surveillance systems could use model-derived risk scores to focus monitoring on patients at elevated genetic or clinical risk for ADRs. Even in drug repurposing, partially predictive models could identify patient subgroups where side effect profiles are more favorable, supporting broader and safer therapeutic applications.

A genetics-based strategy has been proposed aiding in the identification of potential drug targets for a number of human diseases. Recently, our group has created a large catalog of genetic risk factors for hundreds of human phenotypes and mapped those to potentially druggable targets using a Mendelian Randomization (MR)(3) approach. This method leveraged several publicly available data sources as genetic instruments of molecular exposures for gene transcripts or circulating proteins. Our initial analysis was able to identify new targets for hundreds of phenotypes, as well as different repurposing opportunities for previously approved drugs(*6*).

Here we propose to use a similar approach, adding the various mechanisms of actions for existing and to-be-developed drugs, and describe an atlas of drug targets that might lead to different types of ADR. First, we use MR at scale to list the potential safety concerns of targeting all protein-coding genes on 1,708 human phenotypes. We show that our approach is capable of recapitulating hundreds of known ADR for approved drugs. We provide publicly available results where researchers can, for any given protein-coding gene and mechanism of action, retrieve genetic signals that point towards the generation of potential ADR. One advantage of a genomic-led approach is that it does not rely on available selective compounds and as such can be applied early in the drug discovery program to anticipate safety liabilities.

## Results

### A Catalog of Potential Adverse Drug Events

The two-sample MR filtering strategy and the analysis flowchart are represented in Figure 1. We started with 61,156 gene-trait pairs that passed the threshold for MR significance (p-value < 1.59×10-9). From these we mapped the predicted MoA that would lead to an adverse event (e.g., an agonist or inhibitor). Briefly, we used the MR-beta directionality and the safety signal metrics to develop an atlas (Supplementary Table 4), which—given a gene and a pharmacological action of a particular drug—we describe all of the safety concerns for that specific modulation of the target gene.

**Figure 1.**
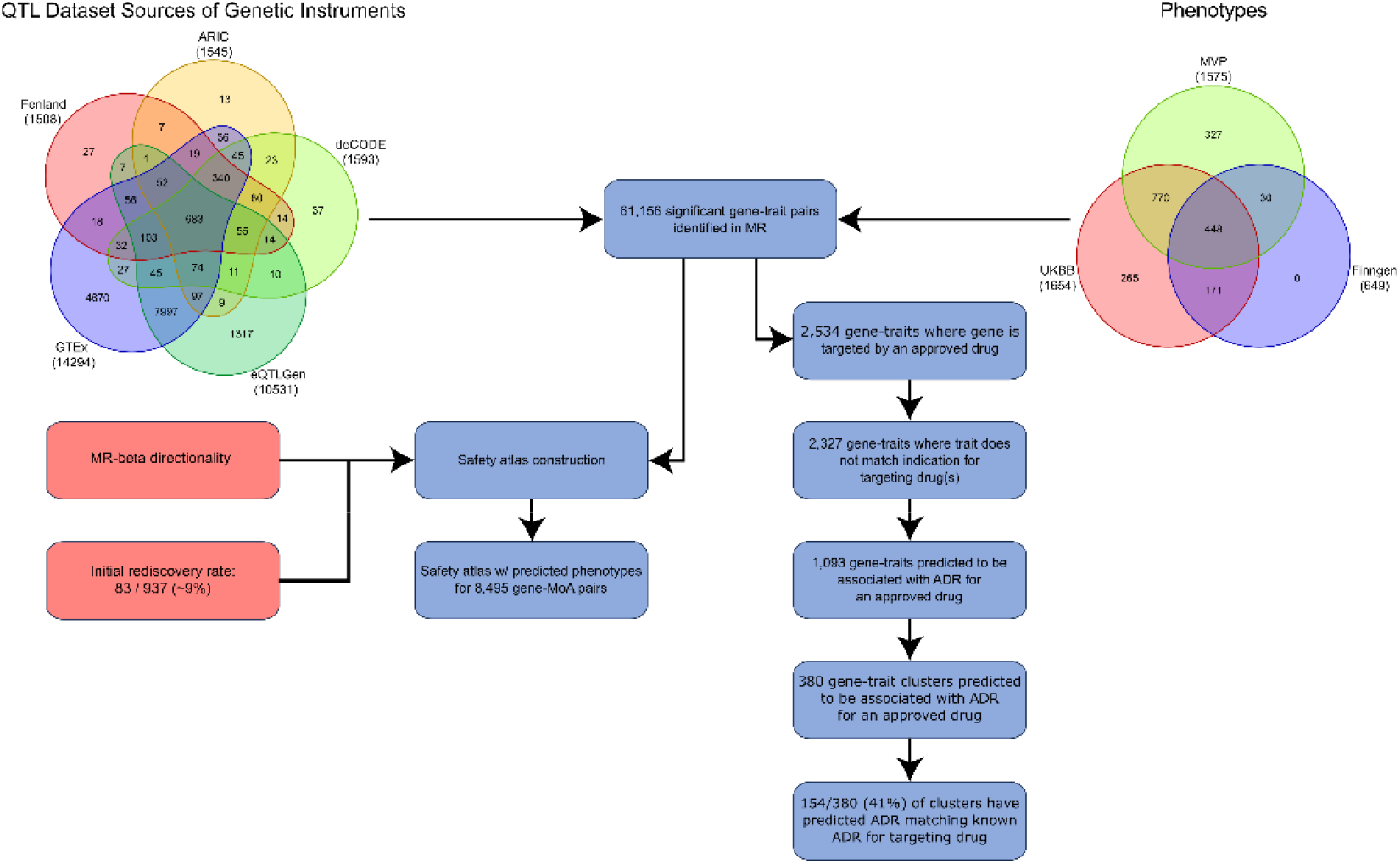
Flowchart of significant gene trait pairs and their corresponding safety events.

Our safety atlas describes predicted phenotypes for 8,495 unique gene-mechanism of action (gene-MoA) pairs. The mean and median number of phenotypes for each unique gene-moa pair was 7.2 and 4.0, respectively, indicating a right skew for the number of unique gene-MoA pairs. The gene HLA-DRB1 was the molecular mechanism predicted to be associated with the highest number of safety concerns (n=176). As expected, regarding those gene-MoA for which there is a chemical compound in development as a treatment, the mean number of phenotypes associated with each gene-MoA was significantly associated with current development phase (Figure 2). Compounds in phase 1 had a median of 14 (mean of 24) safety phenotypes, whereas for approved drugs (phase 4) the median number of associated phenotypes was 4 (mean 6.6), p-value = 4.1e-9.

**Figure 2.**
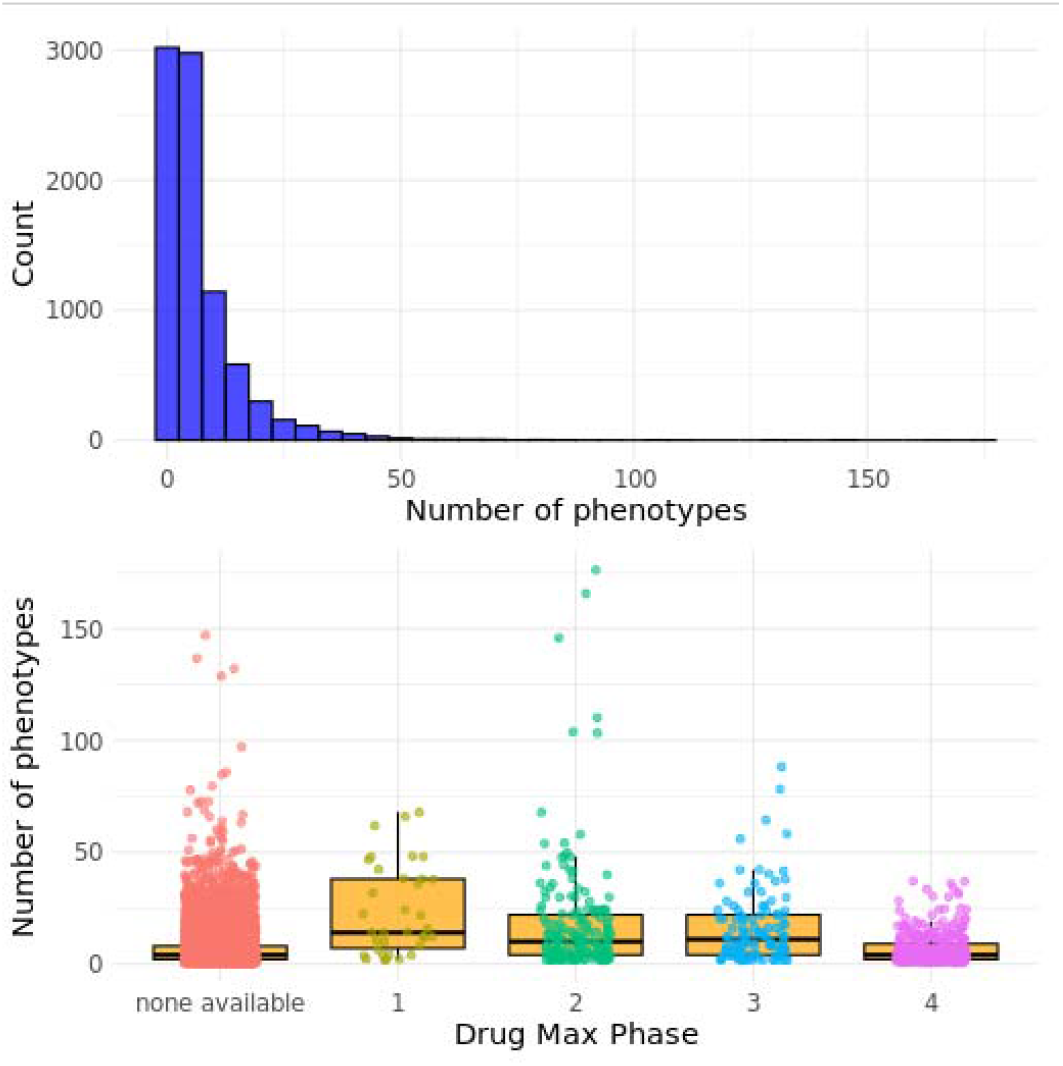
The mean number of phenotypes associated with each current development phase.

### Approximately 40% of genetically identified ADR for genes that target an approved drug have been previously reported in FAERS

Genes that are the target of approved drugs provide another layer of confirmatory evidence for predicted adverse events because these drugs have stood the challenge of post-approval collection of physician and patient refereed adverse drug reactions (ADR). We have specifically analyzed our results in the context of genes and MoA for approved drugs. From the 747 genes targeted by approved drugs, according to the ChEMBL database version 34, there are 249 genes represented in our two-sample MR results. We used a four-step analysis to assess the agreement between an identified ADR and a known ADR as reported by FAERS. Initially, all significant gene-traits in which the gene is targeted by an approved drug were selected. This resulted in 2,534 significant gene-trait pairs (249 different genes and 422 different traits). Next, we excluded those gene-trait pairs in which the trait is an approved drug indication, resulting, after filtering, in 2,327 gene-trait pairs. These were then compared with respect to the MR-predicted directionality of effect (whether increasing or decreasing the exposure to the instrumented molecular phenotype is associated with decreasing or increasing risk of the phenotype). Finally, we contrasted that information with the known mechanism of action of the approved drug targeting the gene. This scheme was repeated for all selected gene-trait pairs. From the 2,327 considered gene-trait pairs our approach identified 1,093 that are predicted to be associated with an ADR for an approved drug. We then used the FAERS database and retrieved all known ADR for all approved drugs targeting the selected genes. This resulted in 205,805 potential ADRs listed in FAERS for the 650 different drugs targeted by the 161 genes identified in this set of results. Since some of the 1,093 traits are very similar, such as “platelet count decreased” and “thrombocytopenia” (Supplementary Table 5), for each gene where several traits were related, we have manually clustered common traits to avoid double counting. This resulted in 380 unique gene-trait clusters where the gene is targeted by a known approved drug. We have systematically mapped all gene-trait clusters to all described ADRs for the approved drugs that target the gene. These gene-trait pairs with known ADRs are represented visually as a network graph in Figure 3. We were able to identify 154 (41%) positive ADR matches among the 380 unique gene-clusters associated with an approved drug (p < 0.001). This result is similar to what we have previously described for the rediscovery of approved drug indications(*6*) among a similar set of approved drugs and is associated with a 1.62-fold enrichment (that is, having MR evidence is associated with higher likelihood of seeing that phenotype listed as AE in FAERS). The fact that 40% of our predicted ADR for approved drugs have been, empirically, previously described for drugs targeting said genes, adds credence to our results.

**Figure 3.**
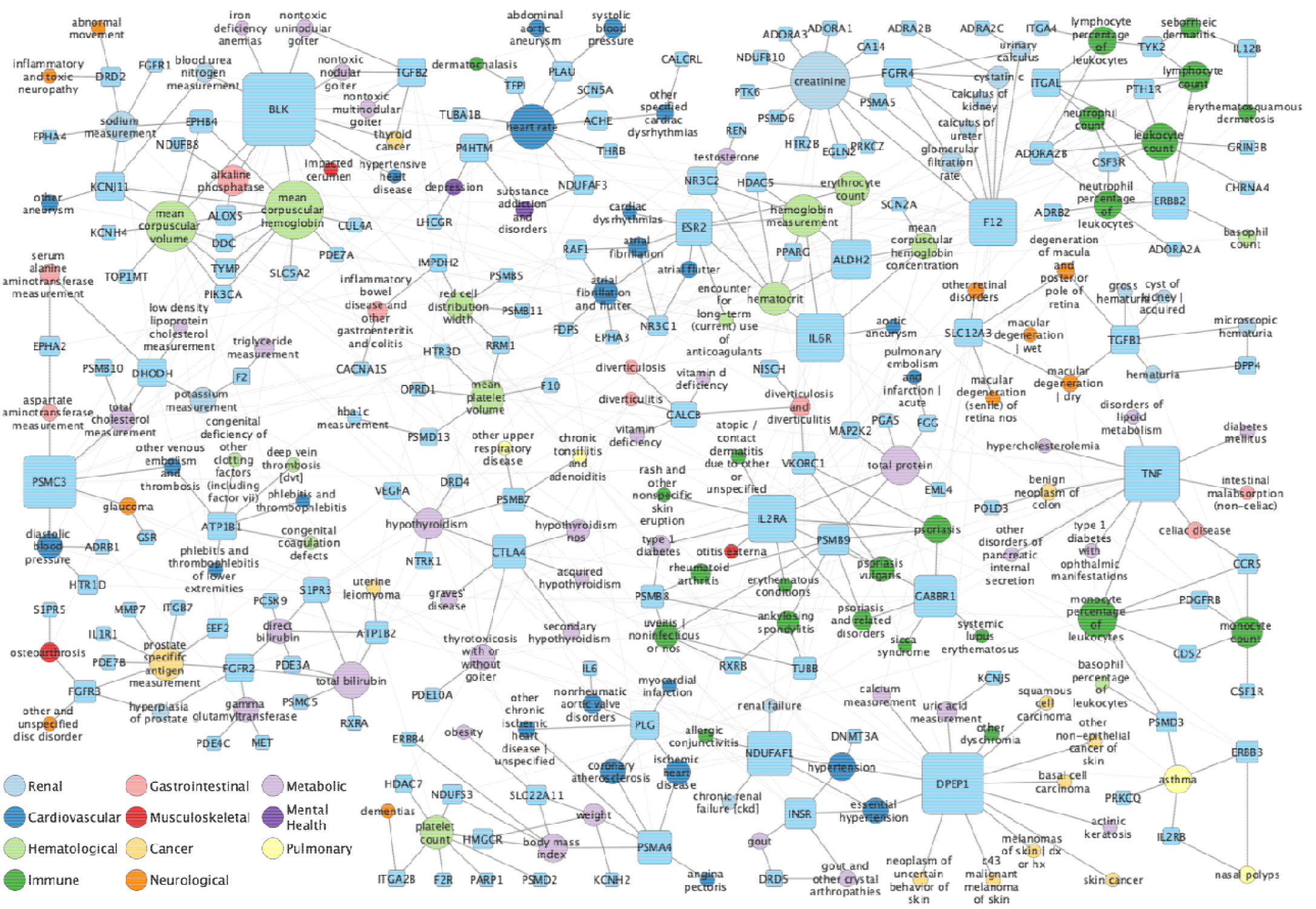
Network visualization of gene-trait pairs with known safety events from FAERS. Nodes represent both genes (square) and traits (circles) with node size scaled by degree, or number of connections. Color represents the high-level category of the trait. Communities were detected using Louvain detection algorithm. For this visualization, we reduce the width of inter-community edges to showcase the communities.

### Genetically Identified ADRs have significantly higher odds of being identified by other sources of genetic information associated with the phenotype

Additionally, we show an expected enrichment of the observed gene-trait pairs that constitutes a significant result from our atlas in relation to other sources of biological annotations, independently, providing support to our approach. We have mapped our safety gene-traits to other existing sources of biological annotations derived from genetic information. These were as follows: the existence of information linking the gene to the same trait in OMIM(*7*), the presence of gene variants causing the phenotype in ClinVar(*8*), the existence of a significant gene burden of damaging rare variants in the gene(*9*) and the presence of the phenotype, and the occurrence of the phenotype when the gene is targeted in a mouse model(*10*). For all tested independent sources of biological support, gene-traits observed to be significant in our approach were also significantly enriched for a positive mapping by these other sources when compared to a set of gene-traits created by shuffling genes and associated phenotypes (Supplementary Table 6, Figure 4).

**Figure 4.**
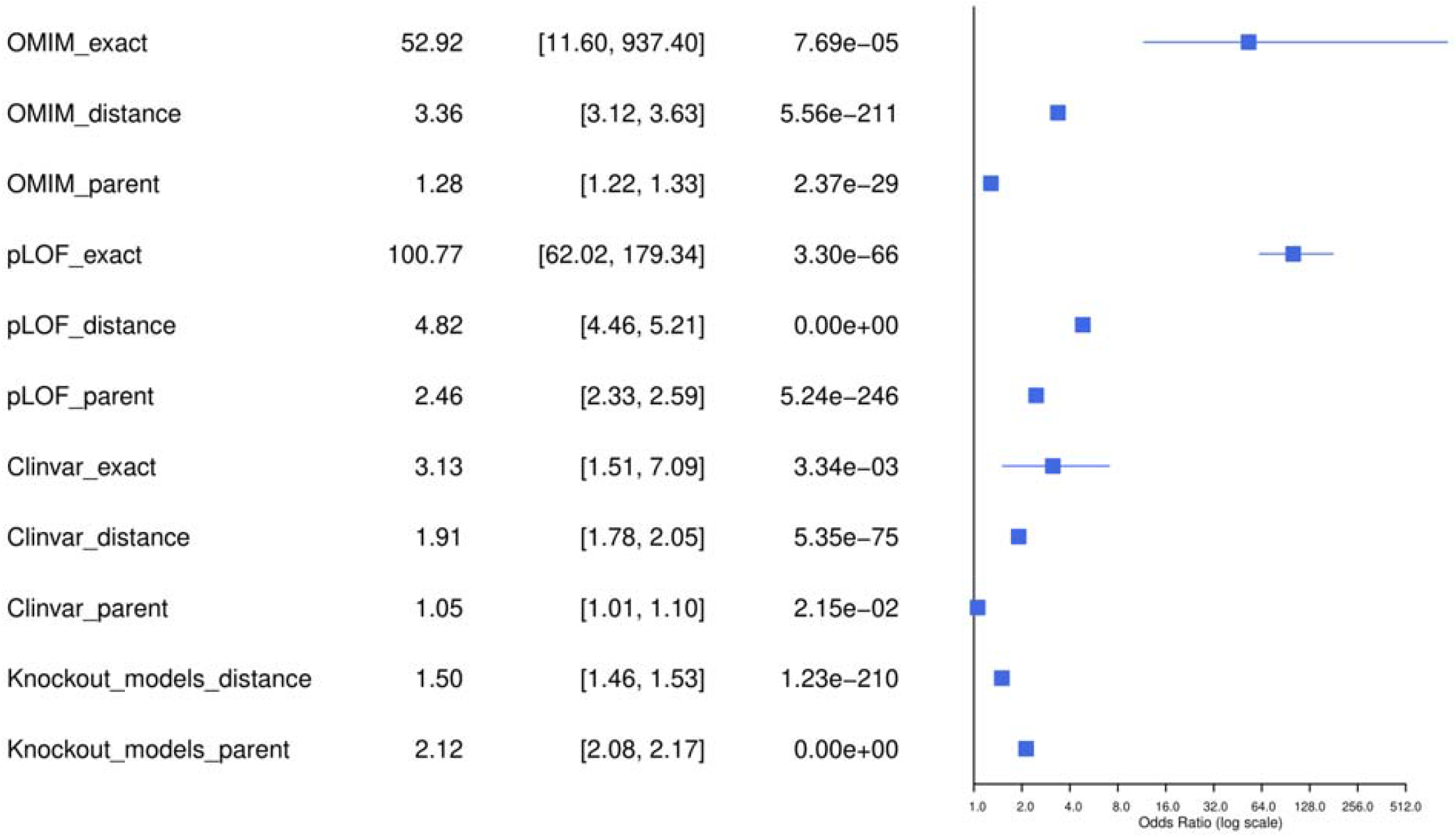
Enrichment of orthogonal sources in significant MR hits.

We did not observe any qualitative difference regarding the enrichment of the used biological annotations when comparing ADR events predicted to be derived from inhibiting the target gene against those derived from activating the target gene (Supplementary Figure 1).

### Genetic factors/patterns associated with genetically predicted ADRs

In order to further explore if we could identify predictive features of a significant gene-ADR MR result we have conducted exploratory analysis annotating our significant gene-ADR pairs for a number of potential predictors. Looking into gene-MoA pairs, we observed 4,142 that were associated with inhibitors and 4,136 that were associated with agonists (and 217 where both an inhibitor and agonist action would lead to an ADR). We observed a median number of 4 (mean 7.3) predicted ADRs for both inhibitors and agonists. In addition, no significant difference in the number of predicted ADR could be associated with the mechanism of action at any stage of drug development (Supplementary Figure 2). Taken together, these results suggest that genetically predicted ADR events are not more commonly associated with agonists than with inhibitors.

Next, we mapped all our genes from significant gene-trait pairs to gnomad conservation metrics (pLI and missense variants z-score). While we did not identify an association between the number of ADR phenotypes and pLI value (a conservation metric for LOF variants), we observed a significant association between the number of ADR phenotypes and the missense variants z-score (rho, 0.04, p-value= 0.0009).

Mapping all the genes from all significant gene-trait pairs to their associated GO(*11, 12*) terms and then deconvoluting this information to understand what GO terms were most associated with the number of genetically predicted ADR events, we were able to initially identify 77 pathways whose modulation is predicted to be associated with a higher number of ADR phenotypes at an FDR-adjusted p-value < 0.0001 (Supplementary Table 7). The vast majority of observed GO terms associated with a higher number of genetically predicted ADR phenotypes were immune-related terms. Since several HLA genes were among the genes associated with the highest number of ADR phenotypes (among the top 8 genes were: HLA-DRB1, HLA-DQA2, HLA-DQB2, HLA-DQB1, HLA-DQA1, HLA-DRB5), we have conducted a sensitivity analysis removing the top 10 genes. Even after removing these genes, we observed 41 significantly enriched GO terms with a preponderance of immune-related pathways such as peptide-antigen binding, acute-phase response, cytokine activity, and positive regulation of T-cell-mediated cytotoxicity (Supplementary Table 8). Interestingly, the clear predominance of immune-related pathways being associated with a higher number of ADR was observed both for positive modulation of identified targets and for negative modulation of identified targets (Supplementary Table 9). These results suggest that our approach has a high sensitivity to predict abnormalities in immune-related traits and that a significant proportion of our identified gene-traits can pleiotropically influence immune and inflammatory ADRs.

### Gene-mechanism of action pairs identified using MR information are significantly enriched in mechanisms of action that fail clinical trials due to safety issues

Finally, we observed a significant association between gene-mechanism of action identified in our efforts and those used in a catalog of clinical trials in which early termination was due to increased safety or toxicity issues. From all drug combinations used in the 269 identified trials with early termination due to safety concerns, we identified 262 unique drugs used. We hypothesize that the molecular mechanism of each of these 262 drugs can potentially be a safety-related molecular mechanism. From these 262 drugs and their primary human gene targets (Supplementary Table 10), we identified 170 distinct gene-mechanism of action pairs on drugs used in failed clinical trials due to safety. Gene-mechanism of action pairs identified in our dataset of failed trials were 1.7-fold more commonly reported in our atlas of genetically mapped safety events than in non-significant gene-mechanism of action pairs (OR = 1.7, 95%CI 1.5 – 1.9, p-value 3.5e-13).

## Discussion

This study presents a systematic approach to leveraging Mendelian randomization (MR) and genetic evidence for identifying adverse drug reactions (ADR) associated with gene-target modulations. By integrating data on gene-trait relationships with pharmacological mechanisms, we developed a comprehensive safety atlas that predicts potential adverse outcomes of therapeutic interventions. This atlas represents a significant advancement in drug safety research, contributing to both drug development and post-market safety assessments.

Our findings align with growing evidence on the utility of human genetics in improving drug safety predictions(*3, 13*). Recent reviews(*3, 13*–*15*) highlight the increasing use of genetic data to predict safety liabilities, especially for novel drug targets lacking robust animal model validation or translational pharmacology data. By identifying 8,495 unique gene-mechanism of action (gene-MoA) pairs and demonstrating their association with varying numbers of phenotypic outcomes, our approach corroborates the concept that genetic proxies can simulate pharmacological modulation effects of target genes.

We demonstrated that gene-trait pairs identified in this atlas are significantly enriched in annotations from databases such as OMIM, ClinVar, and functional mutation burden studies. These findings extend previous work on integrating genomic annotations into drug development pipelines. The enrichment highlights the translational value of combining MR with external genetic datasets to corroborate safety predictions.

### Concordance with Existing Pharmacovigilance Data

The observed 40% overlap between genetically predicted ADRs and those reported in the FAERS database for approved drugs underscores the validity of using MR-based predictions as a pharmacovigilance tool. The significant enrichment of ADR matches (p < 0.001) suggests that genetically derived insights can complement existing data sources, enhancing the accuracy of ADR identification. This result is consistent with previous studies demonstrating the concordance of genetically informed safety signals with clinical observations in post-market surveillance(*13, 16*).

### Pathways and Mechanisms Associated with ADRs

A key observation of this study is the disproportionate representation of immune-related pathways among gene-trait pairs associated with a higher number of genetically predicted ADRs. Pathway analysis revealed significant enrichment for terms such as “peptide-antigen binding” and “cytokine activity,” consistent with earlier findings linking immune modulation to adverse events. Even after sensitivity analyses excluding the top 10 genes, immune pathways remained prominent, reinforcing the hypothesis that genetic variation in immune-related genes frequently underlies ADR risks. Our finding that inhibition of HLA-DRB1 is associated with the highest number of safety concerns (n=176) parallels known clinical challenges with immune-targeting therapies. This observation aligns with prior reports that genetic variants in immune-related loci are strong predictors of therapy-related adverse events, particularly for checkpoint inhibitors. From a practical perspective these findings also emphasize the value of scalable MR analysis in continuous traits that can serve as good proxies of immune-related adverse events, such as lymphocyte counts.

### Clinical Relevance and Future Applications

Our results emphasize the utility of MR as a predictive tool for drug discovery. The observed association between gene-moa pairs and clinical trial terminations due to safety concerns (OR = 1.7, 95% CI: 1.5–1.9, p = 3.5e-13) suggests that genetically-informed safety predictions can guide early-stage decision-making in drug development. This is particularly relevant given the high attrition rates in clinical trials attributed to unanticipated safety issues(*3*). While efficacy remains the leading cause of clinical trial failures, accounting for approximately 57% of Phase II and 45% of Phase III failures, safety concerns are also a significant contributor, responsible for about 17% to 20% of Phase III failures. Conversely, in preclinical stages, safety issues are more prominent, with toxicity being a primary reason for candidate attrition(*17*). Integrating human genetic evidence early in the drug development process can enhance the prediction of both efficacy and safety outcomes.

Interestingly, our initial hypothesis was that agonists would be more frequently associated with ADRs. In other words, that downregulating a gene (decreasing its expression or function) is generally more tolerated than upregulating it (increasing its expression or activity). Physiological systems often feature parallel pathways that can mitigate the effects of partial loss-of-function mutations, providing a level of resilience to inhibitory modulation. However, our findings revealed a comparable distribution of ADRs across mechanisms that increased or decreased the expression of genes, suggesting that this compensatory capacity does not translate into a reduced safety burden for inhibitors.

### Limitations and Future Directions

While our findings demonstrate the utility of Mendelian Randomization (MR) as a predictive tool for drug discovery and highlight the relationship between genetically informed safety signals and clinical trial terminations, several limitations must be acknowledged. The reliance on genetic proxies to infer pharmacological modulation may not capture all nuances of drug action, particularly for polypharmacological agents. Additionally, while the observed enrichments provide strong evidence of validity, functional validation in cellular or animal models remains critical for translating these findings into actionable insights. Future research should explore integrating secondary pharmacology panels and gene expression perturbation data to refine ADR predictions further. Resources such as PharmGWAS(*18*), which systematically combines genetic signatures with drug perturbation profiles, offer promising avenues for expanding the scope of safety assessments.

Although the adverse event data curated by Open Targets from the FDA Adverse Event Reporting System (FAERS) was cleaned and filtered to improve reliability, inherent challenges remain. The FAERS database relies on spontaneous reporting, and its effective population size (that is, the total number of exposed individuals) is unknown, which complicates estimates of absolute or relative risk. Furthermore, despite data cleaning, adverse events that resemble the original indication for which the drug was prescribed often persist in the dataset, making it difficult to distinguish between disease progression, off-target drug effects, or true adverse drug reactions (ADR). These factors may introduce residual confounding and misclassification that could bias our interpretation of gene–mechanism–ADR relationships. Future efforts to integrate pharmacovigilance data with well-annotated exposure denominators, detailed indication information, and prospective validation frameworks will be critical for refining genetically informed safety predictions and improving their translational relevance.

An additional limitation lies in the use and interpretation of clinical trial outcomes. Publicly available trial registries and databases often lack detailed, standardized reporting on the specific reasons for trial failure, particularly when trials are terminated early or fail to meet primary endpoints. Although we focused on trials explicitly classified as terminated for “safety” reasons, classifications can be imprecise or incomplete, with categories such as “business decision” or “strategic realignment” potentially masking underlying safety or efficacy concerns. Moreover, sponsors may underreport unfavorable outcomes to protect proprietary interests, and adverse events leading to discontinuation are not always uniformly adjudicated across different studies. In many cases, failure is multifactorial, with intertwined contributions from efficacy, safety, operational challenges, and commercial factors, making it difficult to assign a single causal attribution. These ambiguities limit the resolution with which we can validate genetically informed safety predictions against trial outcomes and underscore the need for enhanced transparency and structured post-hoc analyses of trial terminations.

## Materials and Methods

### Experimental Design

GWAS summary statistics were obtained from the Million Veteran Program(*19*) (MVP), FinnGen(*20*) R.10, and the UK Biobank (*21, 22*) (UKBB). From UKBB and MVP, we included 1,556 unique Phecodes, 72 biomarkers, 74 questionnaire data, and 6 clinical variables. A total of 1,449 traits (Supplementary Table 1) were harmonized between datasets and meta-analyzed. For meta-analyzed traits, we performed fixed effects inverse-variance weighted meta-analysis using METAL(*23*). For traits where harmonization was not possible (MVP, n=327; UKBB, n=235), we used summary statistics from individual studies. FinnGen on its own was not used. Certain traits with no clear safety concern (for example height) were removed from consideration (n=562) leaving a total of 1,449 traits. Given that the MR framework relies on the expectation that linkage disequilibrium (LD) is the same among genetic studies measuring the association among exposures and outcomes, this study includes samples of European ancestry only. Additional information regarding phenotype harmonization and meta-analysis can be found in “Leveraging Large-Scale Biobanks for Therapeutic Target Discovery”(*6*).

### Instruments for MR

We have used 5 different sources of genetic instruments in the present work. Below we describe the instrument selection criteria for each source.

#### GTEx version 8

Independent cis-eQTLs were identified per gene by performing up to 5 conditional analyses in regions (+/-1 Mb from the transcription start site [TSS] of each gene) using GTEx v8(*24*) individual-level data, additionally iteratively adjusting for the top associated variant if there exists an association reaching a p-value of 1e-4. The primary signal was unconditional for other variants in the region. To identify the independent signals, we considered primary and conditional associations passing a p-value < 5e-8. We then extracted estimates of effect size, corresponding modeled allele, and standard errors from the unconditional association to use in the next steps of MR. This approach was taken for each available GTEx tissue. We used 120,303 genetic markers instrumenting 24,116 genes.

#### eQTLGen

Summary statistics files were downloaded from eQTLGen(*25*). The summary statistics reported SNP-gene associations (<1Mb from the center of the gene and tested in at least 2 cohorts) across 19,250 genes (17,114 in common between GTEx and eQTLGen). We defined instruments using the smallest p-value per gene, with a threshold for inclusion defined as a p-value < XXX. We used 10,669 genetic markers instrumenting 11,175 genes.

#### deCODE

We downloaded the published GWAS of SOMAscan v4 in 35,000 individuals of European ancestry for 4,907 aptamers from deCODE(*26*) (https://www.decode.com/summarydata/). We used 4,775 genetic markers instrumenting 1,624 genes.

#### Fenland

We obtained pQTLs directly from the Fenland study(*27*), a genome-proteome-wide association study among 10,708 participants of European-descent conducted using 10.2 million genetic variants and plasma abundances of 4,775 distinct protein targets (proteins targeted by a least one aptamer) measured using the SOMAscan V4 assay on 4,979 aptamers (4,775 unique protein targets). The unconditional summary statistics from this study are available in an open resource platform (www.omicscience.org). We used 2,881 genetic markers instrumenting 1,510 genes.

#### ARIC

We downloaded the published cis-pQTL GWAS from the Atherosclerosis Risk in Communities (ARIC) study(*28*) (http://nilanjanchatterjeelab.org/pwas/), which contains SOMAscan v4 on 4,657 plasma proteins measured in 7,213 European American individuals. Somamers that mapped to multiple gene targets, lacked a position record in the BioMart database for the target protein-coding gene, or lacked any SNPs in the cis region were excluded from further analysis.

The cis-region is defined as ±500⍰kb of the TSS of the target protein-coding gene in the cis-pQTL analysis. A total of 2,004 significant SOMAmers in the original study were therefore identified as having had at least one cis-pQTL (FDR < 5%) near the gene of the putative protein. We used unconditional estimates from this list of 2,004 cis-pQTLs from the original study. We used 1,612 genetic markers instrumenting 1,594 genes.

### Mendelian Randomization (MR)

Two-sample Mendelian Randomization (MR)(*29*) of each of 16,915 protein-coding genes was performed for all phenotypes using instruments, separately, from five sources of eQTLs and pQTLs. The datasets used for instruments provided summary statistics for the unconditional primary association. For each of the datasets described above, we used the instruments identified by the authors. We extracted the corresponding effect size and standard errors from the unconditional association to use in MR. In order to determine the correct ordering of alleles between the datasets we utilized the harmonise_data() function from the TwoSampleMR package in R. We used the Wald Ratio for instruments with one genetic variant and inverse-variance weighted MR for instruments with multiple genetic variants. We additionally performed MR-Egger for proteins/expression with three or more instruments to be used as a sensitivity analysis. We tested for heterogeneity across variant-level MR estimates, using the Cochrane Q method (mr_heterogeneity option in TwoSampleMR package) and the MR-Egger intercept.

We define genes with significant MR as genes using a Bonferroni correction with p-value ≤ 1.6e-9 for any MR test. This value was obtained by dividing 0.05 by the number of unique gene-trait pairs tested in our study (31,525,236 unique gene-traits initially tested). If a gene was significant in multiple QTL sources, we required the directionality of the beta values to be concordant, otherwise the result was excluded from our list of selected results. If a gene passed in both eQTL and pQTL sources, we only considered the pQTL result.

### Mapping gene results to existing drug data

We extracted drug information for protein targets from ChEMBL (v34)(*30*). For all protein targets, we acquired Ensembl IDs when available using UniProt’s rest API(*31*). For each drug where the information was available in ChEMBL, we identified all indications for those drugs and the clinical phase for that indication. Also added was the mechanism of action (MoA) for the drug and the interaction it has with the target. We further annotated these as positive modulation (activator), negative modulation (inhibitor), or other. Drug information can be found in Supplementary Table 2.

### Adverse drug event data

We used adverse event data reported in the FDA Adverse Event Report System (FAERS) (n = 36,653). This data was acquired from Open Targets(*32*) who processed the FAERS data. Briefly, Open Targets included only reports submitted by healthcare professionals, excluded reports where the outcome was death, and removed events deemed uninformative based on a manually curated exclusion list. We only used drug-ADR that reached statistical significance using the Open Targets approach.

### Clinical trials with early termination due to increased safety concerns

We determined clinical trials that were terminated due to increased safety concerns by systematically filtering trial data. Specifically, we used a previously assembled database of clinical trials (n=17614)(*33*). First, we identified trials flagged as “Terminated,” “Withdrawn,” or “Suspended” in the <overall_status> field to capture all studies that ended earlier than planned. We then reviewed the <why_stopped> field for each record to identify those citing “safety” or “adverse events” as a primary reason for early termination. After excluding duplicates and records in which the stated reason for termination was ambiguous (e.g., “business decision,” “futility,” or “slow enrollment”), 269 trials remained that met our criteria for termination early due to safety concerns. These final 269 trials were included in the subsequent analyses (Supplementary Table 3). The raw trial data, which included various drug and placebo combinations, was parsed to extract unique drug names for each trial arm. A custom R function was employed to clean and parse the drug names from the trial data, ensuring that only drugs with human molecular targets were included, while placebo and other non-drug interventions were excluded. The identified drugs were then mapped to their respective gene-level molecular targets using the existing pharmacological databases DrugBank(*34*) and PubChem(*35*). This mapping allowed us to analyze the molecular mechanisms associated with the drugs used in these trials and their potential contributions to the observed safety concerns. The final dataset of drug-target relationships was compiled and made available for further analysis and integration with other clinical trial data.

## Data Availability

All results are available in Supplementary Materials. Code will be made available upon request.

## Statistical analysis

We compared the median number of identified gene-MoA associations by drug approval phase using a Wilcoxon rank sum test due to the skewed distribution of this variable. We compared the median number of phenotypes associated with negative modulators versus positive modulators using a Kruskal_Wallis rank sum test. The association between the number of ADR and missense or LOF constrain scores was evaluated through a Spearman correlation test.

## Acknowledgements

This research is based on data from the Million Veteran Program, Office of Research and Development, Veterans Health Administration, and was supported by award #MVP000. This publication does not represent the views of the Department of Veteran Affairs or the United States Government.

Full Million Veteran Program acknowledgement can be found in Supplementary File 1.

This research used resources from the Knowledge Discovery Infrastructure at the Oak Ridge National Laboratory, supported by the Office of Science of the U.S. Department of Energy under Contract No. DE-AC05-00OR22725⍰and the Department of Veterans Affairs Office of Information Technology Inter-Agency Agreement with the Department of Energy under IAA No. VA118-16-M-1062.

We would like to acknowledge the time and effort of the study participants and researchers in the Fenland study DOI: 10.22025/2017.10.101.00001; https://www.mrc-epid.cam.ac.uk/research/studies/fenland/.

JCW is funded by the UK Medical Research Council via programme grant MC_UU_00002/18

## Contributions

BRF, JPC, ACP conceived and designed the project.

IAS, BZ, ARL, MK, MG, YT, FH, HI consulted and helped to develop drug related data.

CG, DR, HD curated and assembled QTL data.

BRF, DJG, ACP, KGP, AH developed code for various modules of pipeline.

JCW, AH, KC, JMG, LG, HD provided feedback on experimental design.

IAS, CL, JCW, GMP, GW provided substantive feedback on the manuscript.

SM, TC provided administrative and material support.

RM, LC, NK helped plan and execute the project.

All authors read and approve the manuscript.

## Competing interests

Maya Ghoussaini is a full-time employee at Regeneron Genetics Centre. Maya’s main contributions were while she was an employee of Open Targets. Mohd Karim is a full-time employee at Variant Bio. Mohd’s main contributions were while he was a an employee of Open Targets. JP Casas is a full-time employee at Novartis Institutes for Biomedical Research. JP Casas’ main contributions to the project were while employed at the VA Boston Healthcare System.

### Data and Materials Availability

All results are available in Supplementary Materials. Code will be made available upon request.

