## Supplementary Figures for "A genomic-led strategy to anticipate drug safety effects"

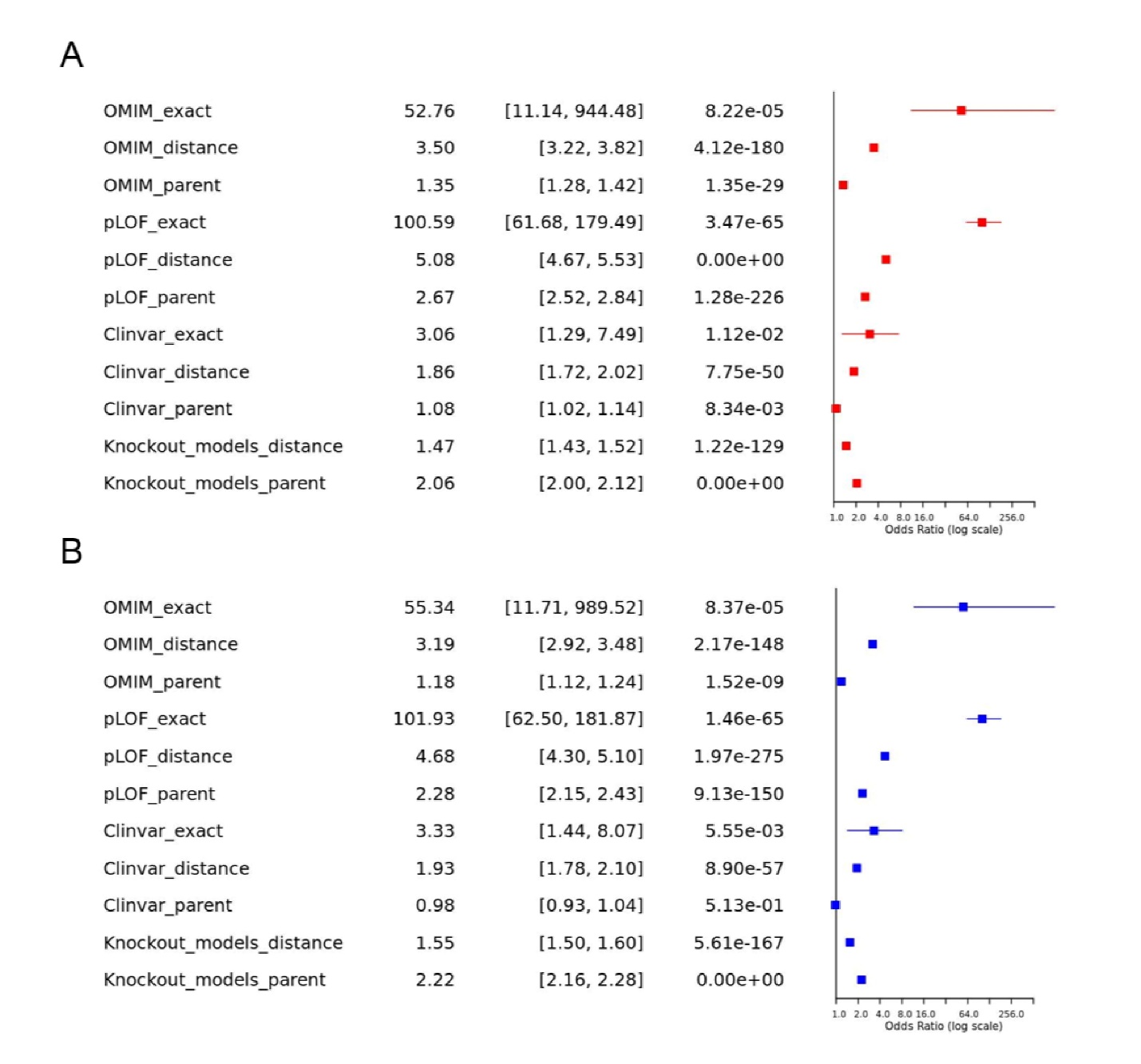

Supplementary Figure 1 – Forest plots of both agonists (A) and inhibitors (B))

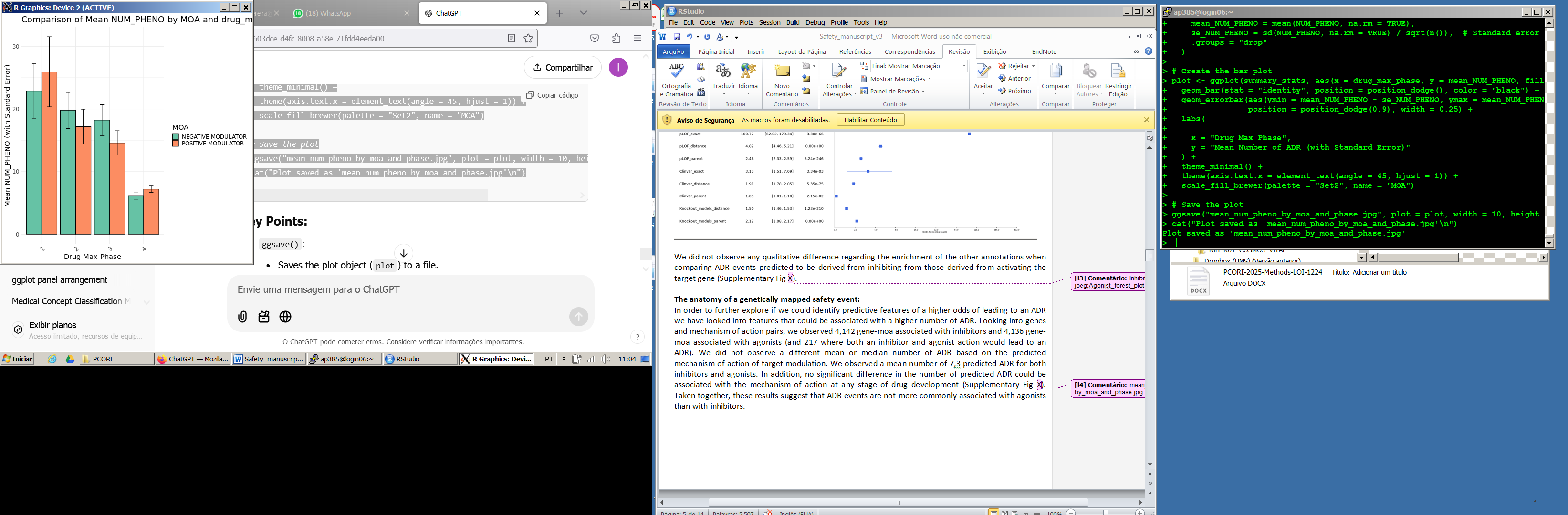

Supplementary Figure 2 – Mean number of phenotypes by both mechanism of action and drug phase.
